# Cervical Spine Assessment Techniques and Neck Strength Profiles of Elite Rugby Union Players Using an Innovative Measurement Approach

**DOI:** 10.1101/2020.06.04.20121988

**Authors:** Lesley McBride, Samuel W Oxford

## Abstract

**Objectives:** Neck injuries and concussion commonly occur in the sport of Rugby Union. This study analyses the current practices of medical staff in assessment of the neck and proposes a new position-specific method using a commercially available setup for the assessment of neck strength.

**Methods:** A survey was employed and distributed to 40 Premiership and Championship Rugby Union clubs. Thirty-eight University students (18 males and 20 females) were tested on two occasions at least 72hrs apart to determine reliability and 131 elite Rugby Union players (forwards n = 82, backs n = 49) were tested on a single occasion. Isometric neck strength was measured using a commercially available rig and digital load cell, for flexion (Flex), extension (Ext), left side flexion (LSF) and right-side flexion (RSF). Peak of three trials for each position was used for statistical analysis.

**Results:** Survey findings indicated no standard practice used in the assessment of the neck across the professional teams. Intra-rater reliability interclass correlation values > 0.924 when using peak neck strength across all directions, thus indicating excellent reliability. Forwards recorded significantly greater absolute values for cervical spine strength across all directions (*p* < 0.01) however, relative values showed no significant difference between players (*p* > 0.05)

**Conclusion:** The results show that the commercially available rig and digital load cell are a reliable tool for the assessment of isometric strength of the cervical spine and provide reference values for healthy males as well as elite Rugby Union forwards and backs.

## INTRODUCTION

Rugby union (rugby) has one of the highest occurrences of reported match and training injuries among contact sports (92/1000 hours,[1, 2] and a growing prevalence of concussion,[2].

The contact situations that characterise rugby carry significant risk of neck damage,[3]. The tackle situation is responsible for the greatest incidence of neck injuries during a match,[4–6]. A recommendation to reduce this injury burden is to strengthen the neck muscles to enable players to dissipate energy from the forces of collisions,[7–11].

Measurements of neck strength do exist in the literature,[8, 12, 13] however, these capacity tests have utilised different modes of assessment,[1]. Previous research,[13] has relied on a custom-built fixed frame which was constructed by the authors in order to generate reliable data sets from the load cells. To date all research into neck strength has also utilised differing start positions thus the results are specific to the research setting but not generalisable to the rugby population. After adjusting for gender and sport,[7] overall neck strength remained a significant predictor of concussion (p = 0.004). Nevertheless homogenous literature on screening and strengthening protocols for the neck is scarce with only four studies being identified where specific neck strengthening protocols were tested in relation to rugby injuries,[11, 14–16]. There is no homogeneous data in the published literature of normative values for neck strength in rugby players.

The aims of this project were to: explore the methods employed in professional rugby to measure neck strength; determine if neck strength can be reliably tested in a simulated contact position using a commercially available rig and gather data on neck strength in professional rugby players

## METHODS

A survey was employed via the online survey platform JISC,[17]. The sample frame was the head of medical services at 40 professional and semi-professional rugby clubs. Questions were designed around key areas incorporating current practice in screening for neck strength and provision of neck strengthening exercise programmes. Questionnaire data was analysed descriptively.

### Reliability Study

Sample size for the estimation of intra-rater reliability of this testing methodology was based on the methods described by Walter and colleagues,[18]. For the values *ρ_o_* = 0.5, *ρ* = 0.8, α = 0.05 and β = 0.02, a total target sample size of 18 participants with a 20 % loss to follow up was necessary. A double-session repeated measures intra-rater reliability study was performed on 38 (18 males and 20 females) recreational healthy adults, recruited by word of mouth. All participants were physically active, had no current neck pain or pathology and were excluded if reported any previous neck injury. Each participant visited the laboratory on two separate occasions, separated by at least 72 hours. Upon arrival the participant’s body mass (kg), height (cm), and neck circumference (cm) were measured. Neck circumference was measured,[8] and each participant completed a standardised warm up which consisted of a static isometric contraction into flexion, extension, right and left side flexion at 80 % maximum voluntary contraction (MVC) against their own hand 3 × 5 seconds whilst standing. The study was approved by the Coventry University Human Research Ethics Committee and before testing participants read and signed an informed consent form.

### Testing procedure

The instruments used in this study included a commercially available ForceFrame™ (Vald Performance, Australia). It comprises an adjustable rig fitted with four independent and adjustable uniaxial load cells. The participants were required to perform three MVC’s in flexion (Flex), extension (Ext), left side flexion (LSF) and right-side flexion (RSF) in a computer-generated randomised order with a 60-second rest between each test. All movements were performed from a standardised quadruped position with the head in contact with the load cell on the ForceFrame ™ and hands were perpendicularly below the shoulder with the scapulae retracted to set the upper quadrant. Elbows remained extended, with the hips and knees at 90 degrees (Figure 1). The load cell was in contact with the frontal bone just above the eyebrows for Flex, the occiput for Ext and the temporal bone just above the superior aspect of the helix of the ear for LSF and RSF. Once the participant had assumed the set position, they performed a familiarisation trial at 50 % of MVC. The participant was instructed to inhale/exhale and then push hard (approximately 3 seconds),[19] verbal encouragement was provided during each MVC,[19, 20]. Three isometric MVC’s were performed for each direction Flex, Ext, LSF and RSF in a randomly determined order.

**Figure 1.**
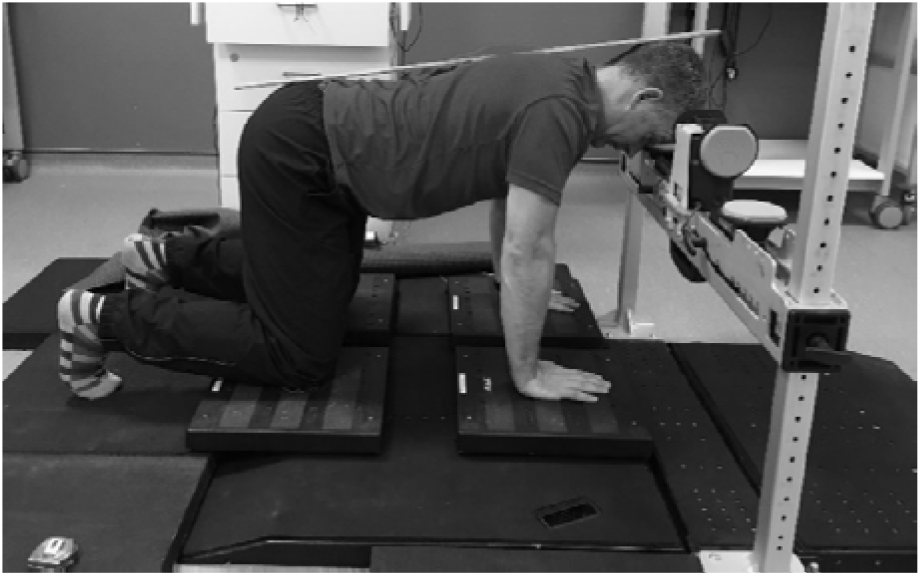
Simulated test position adopted for the assessment of neck strength in the ForceFrame testing rig.

Following the reliability study, the same protocol was used to collect data from 131 professional rugby players from one National team, two Premiership clubs and one Championship club. All testing was carried out indoors at the clubs training facility at least two days post-match and never following a full contact or scrummaging training session. Players were assigned to either the forwards or back unit group based on their playing position.

### Patient and Public Involvement statement

We did not directly include PPI in this study, but patients and public were invited to comment on study design.

### Data Analysis

Force data from the ForceFrame™ were transferred to a personal computer at 50 Hz through a USB connection using custom made software (ForceFrame™, VALD Performance, Australia). The peak and average force for Flex, Ext, LSF and RSF was determined automatically through the ForceFrame™ software and expressed as absolute (N) and relative values (N.kg^−1^). Data analysis was performed with SPSS version (25) (IBM Corp, Armonk, New York, USA) for all statistical analyses. The peak isometric force from each trial was used for statistical analysis. If the peak force recorded was greater than three SD’s of the mean, the trial was excluded from the analysis. To determine the relative reliability of the measures, interclass correlation coefficients (ICCs) were calculated, using the ICC (3,1) for the isometric force values from the three trials for each of the four directions. The ICCs were evaluated using the following criterion measures: poor ICC, 0.50, moderate 0.50, ICC, 0.70, good 0.70, ICC, 0.90, and excellent ICC > 0.90,[21, 22]. Absolute reliability was determined using the standard error of measurement (SEM) by calculating the square root of the within-subject variance [21, 23] while the minimal detectable change (MDC) was determined using the formula MDC = 1.96 * √2 * SEM,[21]

A general linear model of variance was performed to investigate group differences in neck isometric strength for relative and normalised values. The independent variable was group (Males vs Forwards vs Back) and the dependent variables were isometric strength for Flex, Ext, LSF, RSF and total isometric,[24] neck strength as well as the following ratios: F:E and LSF:RSF. Where differences were noted in ANOVA, pairwise comparisons (Bonferroni adjusted) were employed to identify where the significant differences occurred. Effect size for the ANOVA statistics was estimated using partial Eta squared 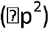 for analysis of variance according to,[25]. Pearson product moment correlation coefficients were determined among anthropometric variables and isometric strength scores. An alpha level of 0.05 was set a priori.

## RESULTS

Descriptive results from the questionnaire are shown in Figure 2.

**Figure 2.**
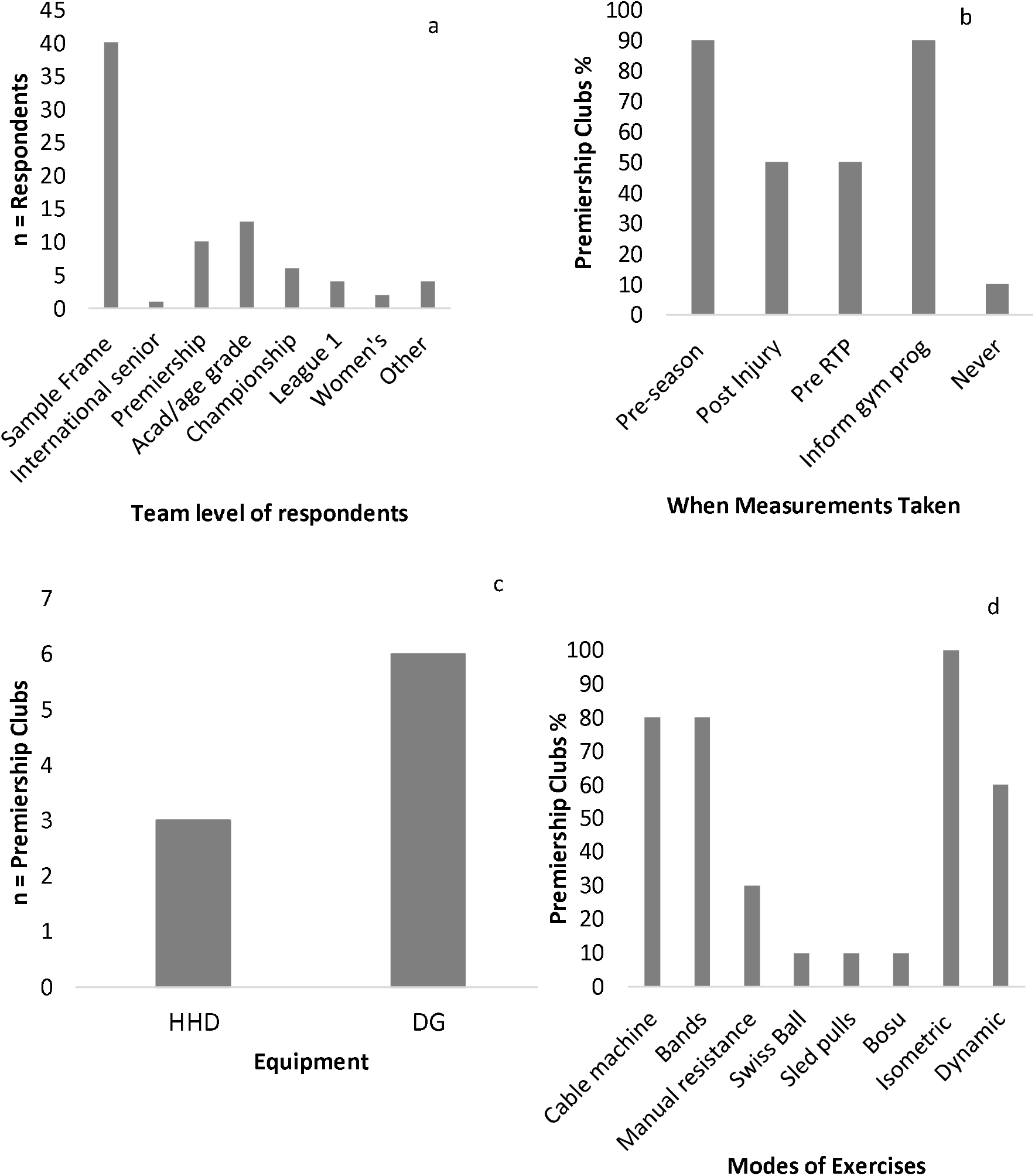
a; The number of respondents from across rugby, b; When premiership rugby clubs measure neck strength, c; The equipment premiership rugby clubs use to measure neck strength and d; The modes of exercise used by premiership rugby clubs.

Abbreviations; HDD = Handheld Dynamometer; DG = Chatillion DG series SS-DG-0210; RTP = Return to Play, Prog = programme

The anthropometric data of the participants is presented in Table 1. There were statistically significant differences across all groups for mass 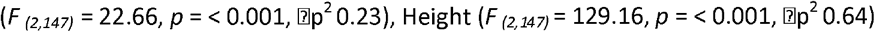, Height 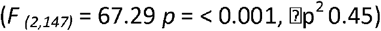 and neck girth 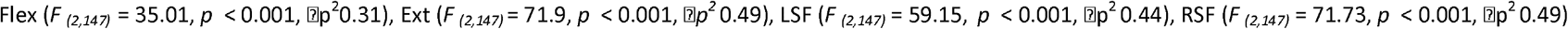 Bonferroni post hoc analysis indicating a significant difference across male and forwards and forwards and backs for mass (*p* = < 0.001), with similar differences reported for height and neck girth between males and backs (Table 1). Descriptive statistics for intra-rater test for each direction are presented in Table 2. The group measure of ICC for Flex, Ext, RSF, and LSF were all excellent and ranged in value from 0.90 to 0.98. The highest SEM was achieved for Ext (43.09 N), indicating the highest level of variability in the four measured directions, whereas RSF was the lowest (5.23 N). When the MDC was compared with the overall mean for each, (Table 2) changes greater than 43.09 N for Ext, 25.67 N for Flex, 25.39 N for LSF, and 25.84 N for RSF would be required to indicate that **a** significant change had occurred in cervical spine strength (Table 2).

**Table 1.** Mean ± SD’s of demographic data

|  | Reliability Group (n=38) |  | Rugby Players (n=131) |  |
| --- | --- | --- | --- | --- |
|  | Males (n=18) | Females (n=20) | Forwards (n=82) | Backs (n=49) |
| Age | 22.74 ± 4.66 | 23.84 ± 8.17 | 24.11 ± 4.42 | 25.38 ± 4.48 |
| Mass (kg) | 86.53 ± 11.58<sup>a</sup> | 64.98 ± 12.92 | 113.75 ± 8.48<sup>a c</sup> | 91.55 ± 8.96<sup>b c</sup> |
| Height (cm) | 180.15 ± 7.90<sup>a b</sup> | 165.80 ± 6.25 | 188.68 ± <sup>a</sup> | 182.45 ± 5.12<sup>b</sup> |
| Neck Girth (cm) | 40.65 ± 2.16<sup>a b</sup> | 33.85 ± 2.20 | 46.33 ± 2.88<sup>a</sup> | 41.98 ± 1.96<sup>a</sup> |

<sup>a</sup> Denotes a significant difference between males and forwards $p < 0.001$
<sup>b</sup> Denotes a significant difference between males and backs $p < 0.001$
<sup>c</sup> Denotes a significant difference between forwards and backs $p < 0.001$

**Table 2.** Intra-rater reliability ICCs, SEMs and MDCs for peak neck strength values.

| Test Position | Test 1 (N) | Test 2 (N) | ICC | SEM (N) | MDC<sub>95</sub> (N) |
| --- | --- | --- | --- | --- | --- |
| Extension | 185.83 ± 85.56 | 192.14 ± 77.84 | 0.960 (0.922 – 0.979) | 9.28 | 25.73 |
| Flexion | 169.56 ± 59.35 | 175.79 ± 56.45 | 0.979 (0.960 – 0.986) | 6.53 | 18.10 |
| Left Side Flexion | 117.53 ± 50.96 | 130.91 ± 47.95 | 0.924 (0.662 – 0.972) | 5.62 | 15.58 |
| Right Side Flexion | 117.62 ± 48.10 | 125.34 ± 44.00 | 0.953 (0.662 – 0.972) | 5.23 | 14.50 |

Abbreviations ICC = interclass correlation coefficient; SEM = standard error of measurement; MDC = minimal detectable change.

There were statistically significant differences across all groups for absolute peak values; 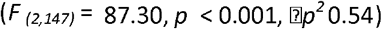 and total isometric strength 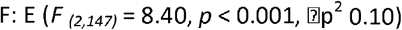 Bonferroni post hoc analysis indicated a significant difference across all groups for Flex, Ext, LSF, RSF and total isometric strength (*p* < 0.001) (Table 3). The ratio values showed a statistically significant difference between groups for 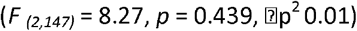 with post hoc analysis showing no difference between backs and males (*p* = > 0.05). There was no significant difference reported for the ratio LSF: RSF 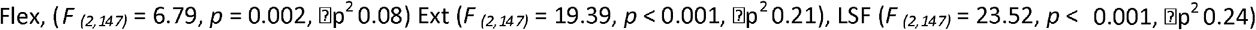 (Table 3). There were statistically significant differences across groups for relative peak force values; 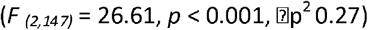. Bonferroni post hoc analysis indicated that there were statistically significant differences between male group and the forwards and backs (p < 0.05), with no significant differences between the forwards and backs (p > 0.05) (Table 3).

**Table 3.** Isometric cervical spine strength values for males, forwards and backs

|  | Control Group (n=18) |  | Forwards (n=80) |  | Backs (n=49) |  |
| --- | --- | --- | --- | --- | --- | --- |
| Test Position | Absolute Peak Force (N) | Relative Peak Force (N.kg<sup>-1</sup>) | Absolute Peak Force (N) | Relative Peak Force (N.kg<sup>-1</sup>) | Absolute Peak Force (N) | Relative Peak Force (N.kg<sup>-1</sup>) |
| Flexion | 218.75 ± 40.78<sup>a b</sup> | 2.57 ± 0.58<sup>a b</sup> | 338.37 ± 71.91<sup>a c</sup> | 2.98 ± 0.60<sup>a</sup> | 284.33 ± 45.94<sup>b c</sup> | 3.11 ± 0.47<sup>b</sup> |
| Extension | 250.55 ± 71.35<sup>a b</sup> | 2.92 ± 0.80<sup>a b</sup> | 443.45 ± 76.85<sup>a c</sup> | 3.90 ± 0.59<sup>a</sup> | 335.20 ± 66.08<sup>b c</sup> | 3.66 ± 0.61<sup>b</sup> |
| F:E | 0.93 ± 0.28<sup>a b</sup> |  | 0.77 ± 0.15<sup>a</sup> |  | 0.87 ± 0.17<sup>b</sup> |  |
| LSF | 151.65 ± 50.65<sup>a b</sup> | 1.72 ± 0.61<sup>a b</sup> | 305.18 ± 72.06<sup>a c</sup> | 2.68 ± 0.59<sup>a</sup> | 228.76 ± 49.15<sup>b c</sup> | 2.50 ± 0.48<sup>b</sup> |
| RSF | 150.57 ± 45.03<sup>a b</sup> | 1.76 ± 0.54<sup>a b</sup> | 309.48 ± 69.22<sup>a c</sup> | 2.72 ± 0.57<sup>a</sup> | 226.94 ± 41.43<sup>b c</sup> | 2.49 ± 0.42<sup>b</sup> |
| LSF:RSF | 1.01 ± 0.16 |  | 1.02 ± 0.14 |  | 1.01 ± 0.12 |  |
| Total Isometric Strength | 771.52 ± 180.09<sup>a b</sup> | 8.98 ± 2.19<sup>a b</sup> | 1396.48 ± 238.39<sup>a c</sup> | 12.28 ± 1.88<sup>a</sup> | 1075.23 ± 163.90<sup>b c</sup> | 11.79 ± 1.48<sup>b</sup> |

<sup>a</sup> Denotes a significant difference between males and forwards $p < 0.001$
<sup>b</sup> Denotes a significant difference between males and backs $p < 0.001$
<sup>c</sup> Denotes a significant difference between forwards and backs $p < 0.001$

## DISCUSSION

This study set out to establish the current practices being used for the assessment and management of the neck in professional rugby clubs. This has not previously been reported in the literature thus making this a novel piece of research. The importance of a strong neck for rugby had been previously stated but not yet defined,[26]. The questionnaire revealed that no standardised method of neck strength assessment is being adopted (Figure 2, c) and a variety of modes of strengthening are utilised (Figure 2, d). Neck strength has been shown to change throughout a season therefore measuring it only once pre-season (Figure 2, b) requires consideration, [27]. The most popular mode of exercise was isometric (Figure 2, d) despite a lack of evidence supporting an improvement in dynamic strength,[28]. The initial survey identified the need for a valid and reliable accessible method for the measurements of neck strength and the development of a consensus of what constitutes a strong neck in rugby.

This study demonstrated that our novel isometric neck strength measurement protocol, simulating a rugby contact posture, achieved excellent reliability results in line with those previously reported,[29] for abductor strength using the commercially available ForceFrame™ and digital force gauge (Vald Performance Australia). The study incorporated a standardised neck strength testing protocol and frame setup. The protocol can easily be replicated and provided intra-rater reliability with ICC values ranging from (0.924 –0.979) when using peak cervical strength values. The intra-rater reliability results are similar to other neck strength studies that have utilised a custom made rig and commercially available digital force gauges 0.901–0.972 and 0.958 – 0.990 respectively, [30]. Previous studies that have used various techniques and devices have also reported excellent reliability for intra-rater ICC’s ranging between 0.930 –0.990 [31] and 0.940–0.990,[32].

Most previous studies investigating neck strength have conducted testing in a seated position with the torso restrained, [33, 34]. Trunk stabilisation is reported, [13] to be essential for reliable and valid neck strength measurements, [34], however trunk stabilisation limits construct validity and the functional relevance of the strength measures, [13]. In this study the trunk was stabilised through the retraction of the scapulae and the horizontal body position with the elbow extended perpendicular to the shoulders and 90° hip and knee flexion. A previous study,[13] used a similar body position, but with the trunk supported on a bench. The absolute values reported for the control group (Table 3) in the current study were 23 – 83 N stronger than those reported in a similar population,[19] but regardless of the strength differences the patterns reported for each movement direction were similar.

To determine the clinical significance of the obtained differences absolute reliability scores (SEM) were calculated for each direction (Ext: 9.28 N, Flex: 6.53 N, RSF: 5.23 N and LSF: 5.62 N), indicating the standard deviation expected in peak force values when repeatedly testing an individual,[35]. Previous work examining SEM values for cervical musculature in a simulated contact position,[19] reported values of below 19 N indicated small variation in these values. This study reported values considerably below these values. The reporting of the MDC has particular importance when assessing changes in interventions or for day to day clinical decisions. In this study for the control group a change greater than 5.73 N for Ext, 18.10 N for Flex, 15.58 N for LSF and 14.50 N for RSF classed as a MDC.

Rugby players were assigned to a forwards and back unit based on playing position. The results of this study indicate that the isometric neck strength profile of the rugby players significantly (*p* < 0.001) exceeds that of the general population (Table 3) in line with previous research, [12]. Considering the physical nature of the sport this is not surprising but reflects the importance for rugby players to possess strength in this region of the body. A significant difference (p < 0.001) was observed in the absolute isometric neck strength values across all measured planes of motion of the forwards and backs (Table 3). The forwards are expected to have significantly greater extension strength due to the positional demands of scrummaging, rucking and mauling and the angle of force application in the vertical plane requiring stability achieved through strong neck extension, [34]. There is also large transverse and horizontal force in the binding between the front row and second row players,[34].

The values for neck extension in this study for forwards and backs (443.45 ±76.85 N vs 335.20 ± 66.08 N), are lower than previous values reported (637.10 ± 75.15 N vs 537.87 ± 82.25 N) on rugby players,[12]. The same profile of results was similar for the other movement directions. These results may be lower due to the differences in testing set-up: in the previous study participants were seated and required to resist a force applied by the tester whereas this study measured the athlete’s capacity to generate force which would provide a more suitable clinical assessment and safer test for the neck, allowing for earlier testing in the recovery process post injury than the aforementioned test,[12].

The ratio values of the forwards and backs for F:E (0.77± 0.15 vs 0.87 ± 0.17) were also higher than the previous values reported on rugby players, [12] perhaps due to the increased importance placed on neck strength and its relationship with concussion, which is the most frequently occurring injury in the professional game,[36]. Further research is required to understand the importance of the ratio of F:E and LSF:RSF rather than simply neck strength. A previous study,[4] highlighted that an average of 456.8 contact situations occur per game with the tackle being the most common (2212 events per game) followed by rucks (142 events per game). The tackle is the most injurious event that takes place in rugby accounting for between 40 – 60 % of all match injuries,[36, 37] with head injuries occurring most frequently during front on and high tackles. Increases in neck strength are related to reductions in head accelerations during the tackle situation in rugby union players, [38] who concluded that increasing neck strength may reduce concussion risk in contact sports. The ratio values for LSF:RSF (1.02 ± 0.14 vs 1.01 ± 0.12) are in line with those previous published,[12]. Greater neck strength and anticipatory muscle activation are associated with an attenuated response to impulsive force(s) acting on the athletes head, [9, 39]. Associated head impacts in rugby occur in all directions and there is evidence to suggest that coronal plane lateral flexion may be more injurious than head motion primarily involving sagittal plane flexion and extension,[40]. The neck musculature plays a protective role in controlling the dynamic head response to trauma during contact situations and co-contraction should increase the necks ability to generate stiffness and therefore to resist head motion due to external forces, [9].

Low neck strength is a potentially a modifiable risk factor that may contribute to the elevated risk of concussion, [9]. Stronger muscles are not only capable of generating greater absolute force values, but have a greater cross-sectional area, greater tensile stiffness at a given activation level and generate torque more rapidly than weaker muscles,[41]. Having lower levels of strength may be a disadvantage regarding controlling the head’s response during an impact because of lower tensile stiffness to resist cervical muscle stretch, [9]. When the cervical spine strength scores are considered as relative values of body mass (kg) (Table 3) then the results show that there is no significant difference between the forwards and the backs (p > 0.05). Indicating the importance of implementing a monitoring programme for all players and therefore determining positional specific values.

A limitation of the current study is that data was collected during the first quarter of the season. Ideally the players would be measured pre-season and then at intervals during the season. Future research should consider investigating the differences in each playing position rather than considering forwards and backs as independent groups.

## CONCLUSION

This new protocol and strength testing rig (ForceFrame™) can be used to identify individuals that may be at greater risk of injury due to neck strength limitations and can be used to inform gym programmes. This new protocol allows a player centred approach to the assessment and management of the neck throughout a season. Incorporating this measurement protocol in a club setting would allow for the assessment of the association between neck strength and concussion.

## Data Availability

The data that support the findings of this study are available from the corresponding author, upon reasonable request.

## ACKNOWLEDGEMENTS

The ForceFrame setup was loaned by Vald Peformance for the duration of the study. The company were not involved in the design, data analysis or interpretation of the results and neither author has any commercial interests in the company.

## References

1 Hrysomallis C. Neck muscular strength, training, performance and sport injury risk: a review, Sports Medicine 2016;46:1111–24.

2 West S. England Professional Rugby Injury Surveillance Project: 2017/18. 2018, Rugby Football Union (RFU):l-60.

3 Küster D, Gibson A, Abboud R, et al. Mechanisms of cervical spine injury in rugby union: a systematic review of the literature, Br J Sports Med 2012;46:550–4.

4 Fuller CW, Brooks JH, Cancea RJ, et al. Contact events in rugby union and their propensity to cause injury, Br J Sports Med 2007;41:862–7.

5 Tucker R, Raftery M, Kemp S, et al. Risk factors for head injury events in professional rugby union: a video analysis of 464 head injury events to inform proposed injury prevention strategies, Br J Sports Med 2017;51:1152–7.

6 Trewartha G, Preatoni E, England ME, et al. Injury and biomechanical perspectives on the rugby scrum: a review of the literature, Br J Sports Med 2015;49:425–33.

7 Collins CL, Fletcher EN, Fields SK, et al. Neck strength: a protective factor reducing risk for concussion in high school sports, The journal of primary prevention 2014;35:309–19.

8 Hamilton DF, Gatherer D, Robson J, et al. Comparative cervical profiles of adult and under-18 front-row rugby players: implications for playing policy, BMJ open 2014; 4:e004975.

9 Eckner JT, Oh YK, Joshi MS, et al. Effect of neck muscle strength and anticipatory cervical muscle activation on the kinematic response of the head to impulsive loads, Am J Sports Med 2014;42:566–76.

10 Lisman P, Signorile JF, Del Rossi G, et al. Cervical Strength Training Does Not Enhance Dynamic Stabilization of Head and Neck During Football Tackling. 2010;42:679.

11 Naish R, Burnett A, Burrows S, et al. Can a specific neck strengthening program decrease cervical spine injuries in a men’s professional rugby union team? A retrospective analysis, Journal of sports science & medicine 2013;12:542.

12 Geary K, Green BS, Delahunt E. Intrarater reliability of neck strength measurement of rugby union players using a handheld dynamometer, J Manipulative Physiol Ther 2013;36:444–9.

13 Salmon DM, Handcock PJ, Sullivan SJ, et al. Reliability of repeated isometric neck strength and endurance testing in a simulated contact posture, The Journal of Strength & Conditioning Research 2015;29:637–46.

14 Geary K, Green BS, Delahunt E. Effects of neck strength training on isometric neck strength in rugby union players, Clinical Journal of Sport Medicine 2014;24:502–8.

15 Barrett MD, Mcloughlin TF, Gallagher KR, et al. Effectiveness of a tailored neck training program on neck strength, movement, and fatigue in under-19 male rugby players: a randomized controlled pilot study, Open access journal of sports medicine 2015;6:137.

16 Maconi F, Venturelli M, Limonta E, et al. Effects of a 12-week neck muscles training on muscle function and perceived level of muscle soreness in amateur rugby players, Sport Sciences for Health 2016;12:443–52.

17 Bristol Online Survey. Online survey tool, 2016.

18 Walter SD, Eliasziw M, Donner A. Sample size and optimal designs for reliability studies. Stat Med 1998;17:101–10.

19 Salmon DM, Handcock PJ, Sullivan SJ, et al. Can neck strength be measured using a single maximal contraction in a simulated contact position? The Journal of Strength & Conditioning Research 2018;32:2166–73.

20 Strimpakos N. The assessment of the cervical spine. Part 2: strength and endurance/fatigue, J Bodywork Movement Ther 2011;15:417–30.

21 Hopkins WG. Measures of reliability in sports medicine and science. Sports medicine 2000;30:1–15.

22 Roebroeck ME, Harlaar J, Lankhorst GJ. The application of generalizability theory to reliability assessment: an illustration using isometric force measurements. Phys Ther 1993;73:386–95.

23 Beckerman H, Roebroeck ME, Lankhorst GJ, et al. Smallest real difference, a link between reproducibility and responsiveness. Quality of Life Research 2001;10:571–8.

24 Dvir Z, Prushansky T. Cervical muscles strength testing: methods and clinical implications. J Manipulative Physiol Ther 2008;31:518–24.

25 Ferguson CJ. An effect size primer: a guide for clinicians and researchers. 2016.

26 Hamilton DF, Gatherer D. Cervical isometric strength and range of motion of elite rugby union players: a cohort study, BMC Sports Science, Medicine and Rehabilitation 2014;6:32.

27 Danielle S, Pinfold J, Sullivan SJ, et al. Descriptive analysis of head impact kinetics in a simulated rugby union tackle: preliminary findings, Br J Sports Med 2017;51:A4.

28 Cazzola D, Holsgrove TP, Preatoni E, et al. Cervical spine injuries: A whole-body musculoskeletal model for the analysis of spinal loading, PLoS One 2017;12.

29 Ryan S, Kempton T, Pacecca E, et al. Measurement properties of an adductor strength-assessment system in professional australian footballers. International journal of sports physiology and performance 2019;14:256–9.

30 Hall T, Morissette MP, Cordingley D, et al. Reliability of a Novel Technique for the Measurement of Neck Strength, International Journal of Athletic Therapy and Training 2017;22:43–50.

31 Strimpakos N, Sakellari V, Gioftsos G, et al. Intratester and intertester reliability of neck isometric dynamometry, Arch Phys Med Rehabil 2004;85:1309–16.

32 Almosnino S, Pelland L, Stevenson JM. Retest reliability of force-time variables of neck muscles under isometric conditions, Journal of athletic training 2010;45:453–8.

33 Dvir Z, Prushansky T. Cervical muscles strength testing: methods and clinical implications. J Manipulative Physiol Ther 2008;31:518–24.

34 Olivier PE, Du Toit DE. Isokinetic neck strength profile of senior elite rugby union players, Journal of Science and Medicine in Sport 2008;11:96–105.

35 Edmondston SJ, Wallumrød ME, MacLéid F, et al. Reliability of isometric muscle endurance tests in subjects with postural neck pain. J Manipulative Physiol Ther 2008;31:348–54.

36 Kemp S, West S, Brooks J, et al. England Professional Rugby Injury Surveillance Project: 2017–18 Report, Union, RF (Ed.) 2019.

37 Brooks JH, Fuller CW, Kemp S, et al. Epidemiology of injuries in English professional rugby union: part 1 match injuries, Br J Sports Med 2005;39:757–66.

38 Dempsey AR, Fairchild TJ, Appleby BB. The relationship between neck strength and head accelerations in a rugby tackle. 2015.

39 Tierney RT, Sitler MR, Swanik CB, et al. Gender differences in head-neck segment dynamic stabilization during head acceleration, Medicine & Science in Sports & Exercise 2005;37:272–9.

40 Patton DA, Mcintosh AS, Kleiven S. The biomechanical determinants of concussion: finite element simulations to investigate tissue-level predictors of injury during sporting impacts to the unprotected head, Journal of applied biomechanics 2015;31:264–8.

41 Thelen DG, Ashton-Miller JA, Schultz AB, et al. Do neural factors underlie age differences in rapid ankle torque development? J Am Geriatr Soc 1996;44:804–8.

